# Predicting and simulating effects of PEEP changes with machine learning

**DOI:** 10.1101/2021.01.28.21250212

**Authors:** Claas Strodthoff, Inéz Frerichs, Norbert Weiler, Björn Bergh

**Affiliations:** Department of Anaesthesiology and Intensive Care Medicine, University Medical Centre Schleswig-Holstein, Campus Kiel, Kiel, Germany; Department of Medical Informatics, University Medical Centre Schleswig-Holstein, Campus Kiel, Kiel, Germany

**Keywords:** MIMIC-III, eICU, PEEP, Ventilation, Deep Learning, artificial neural networks

## Abstract

**Background/Objectives:** Choosing ventilator settings, especially positive end-expiratory pressure (PEEP), is a very common and non-trivial task in intensive care units (ICUs). Established solutions to this problem are either poorly individualised or come with high costs in terms of used material or time. We propose a novel method relying on machine learning utilising only already routinely measured data.

**Methods:** Using the MIMIC-III (with over 60000 ICU stays) and eICU databases (with over 200000 ICU stays) we built a deep learning model that predicts relevant success parameters of ventilation (oxygenation, carbon dioxide elimination and respiratory mechanics). We compare a random forest, individual neural networks and a multi-tasking neural network. Our final model also allows to simulate the expected effects of PEEP changes.

**Results:** The model predicts arterial partial pressures of oxygen and carbon dioxide and respiratory system compliance 30 minutes into the future with mean absolute percentage errors of about 22 %, 10 % and 11 %, respectively.

**Conclusions:** The deep learning approach to ventilation optimisation is promising and comes with low cost compared to other approaches.

## I. Introduction

Mechanical ventilation is one of the central purposes of intensive care units (ICU). Selection of ventilator parameters is being performed countless times every day worldwide. One of the most controversial settings is the positive end-expiratory pressure (PEEP). Large clinical studies showed the importance of PEEP selection for patient mortality, although not in a consistent way [1], [2]. The least one can conclude from these studies is that setting an individually adequate PEEP is an important and difficult task.

There is a range of established methods for PEEP optimisation. PEEP can be set as a function of the required oxygen fraction [3] or chosen to optimise compliance following a PEEP trial [4]. More sophisticated methods make use of additional medical equipment such as an esophageal pressure probe [5] or electrical impedance tomography [6], [7]. While these are very promising approaches which offer new insight into the topic, their use is associated with extra material cost and it requires extra effort of trained personnel. Probably the most common method to optimise PEEP relies on personal clinical experience as well as trial and error, which is hard to evaluate objectively. All in all, there is no consensus about the best approach to optimise PEEP.

In this paper we present an alternative tool which does not have the aforementioned drawbacks of lacking individualisation or high cost. Taking advantage of the advances in machine learning, in particular deep learning, as well as the current development to record and save more and more data in the ICU setting, we developed a deep learning model to help with this decision. The aim of mechanical ventilation is to deliver sufficient gas exchange while at the same time causing as little harm as possible to the patient. Therefore, with this approach we offer the clinician real-time information and also simulations for relevant target variables of oxygenation, carbon dioxide elimination and respiratory system mechanics.

Machine learning-based clinical decision support systems become increasingly common in the ICU [8]. With respect to this work, some publications are especially important: Similar to our study, Ghazal et al. tried to predict oxygen saturation in infants after a change in ventilator settings [9]. The authors ultimately did not reach satisfactory accuracy, supposedly because of insufficient data. Relying on classical machine learning techniques, Luo et al. predicted laboratory results from previously known other laboratory results [10], however, without incorporating other measurement data. Komorowski et al. aimed to find the best treatment regime for the circulatory system (catecholamines versus intravenous fluids) in sepsis [11]. They used a reinforcement learning approach, which is considerably more complicated than the supervised learning approach used in this work but offers the possibility to consider the treatments’ long-term effects like mortality more easily.

## II. Methods

### A. Data Sources

1. *MIMIC-III dataset:* In the field of intensive care databases for machine learning there is one database that stands out in its size, quality and frequency of use in machine learning publications. Medical Information Mart for Intensive Care III (MIMIC-III) is an openly accessible database of over 40000 patients that were treated in the ICU of the Beth Israel Deaconess Medical Center in Boston between 2001 and 2012 [12]. The database includes demographic data, vital signs, laboratory results, interventions, medication, notes of the medical personnel, radiologic findings and mortality data of over 60000 ICU stays. We used this database as a training and test (10%) set.
2. *eICU dataset:* The eICU Collaborative Research Database holds data associated with over 200000 patient stays recorded in numerous ICUs in the United States, made available by Philips Healthcare in partnership with the MIT Laboratory for Computational Physiology [13]. As far as the contents are concerned, this database is very similar to the MIMIC-III database with the advantage of being more diverse because of its multiple origins. In this work we used this second database as an additional independent test set which the model never sees during training.

For both databases we only used data from ventilated patients at least 16 years old.

### B. Data Preprocessing

We started with all time points where the desired target variable has been measured (ground truth). For each of these events, we collected the desired input variables according to a predefined template from the minutes and hours before (see table I). The used items have different characteristics: items can either be set (F_i_O_2_, PEEP, norepinephrine rate) or measured (S_p_O_2_, temperature, mean arterial pressure); they can be timed (S_p_O_2_, F_i_O_2_) or constant over one ICU stay (age, height); they can be continuous (S_p_O_2_, temperature) or categorial (ventilator mode, sex). From these items we always took the latest measurement that was available before the time of measurement of the target variable. In order to include older measurements for some items, we discarded the latest *n* minutes before selecting a measurement (denoted as “item name *[n]*”). Whenever columns contained missing values, an extra boolean column indicating the presence/absence of values was added and the missing values were imputed as the mean. We performed only minimal data curation: removal of impossible values and conversion of units or representations to be identical for one variable in case they varied considerably between but also within one database(s) (temperature, height, weight, fractions vs. percentages, ventilator mode names).

**TABLE I:**
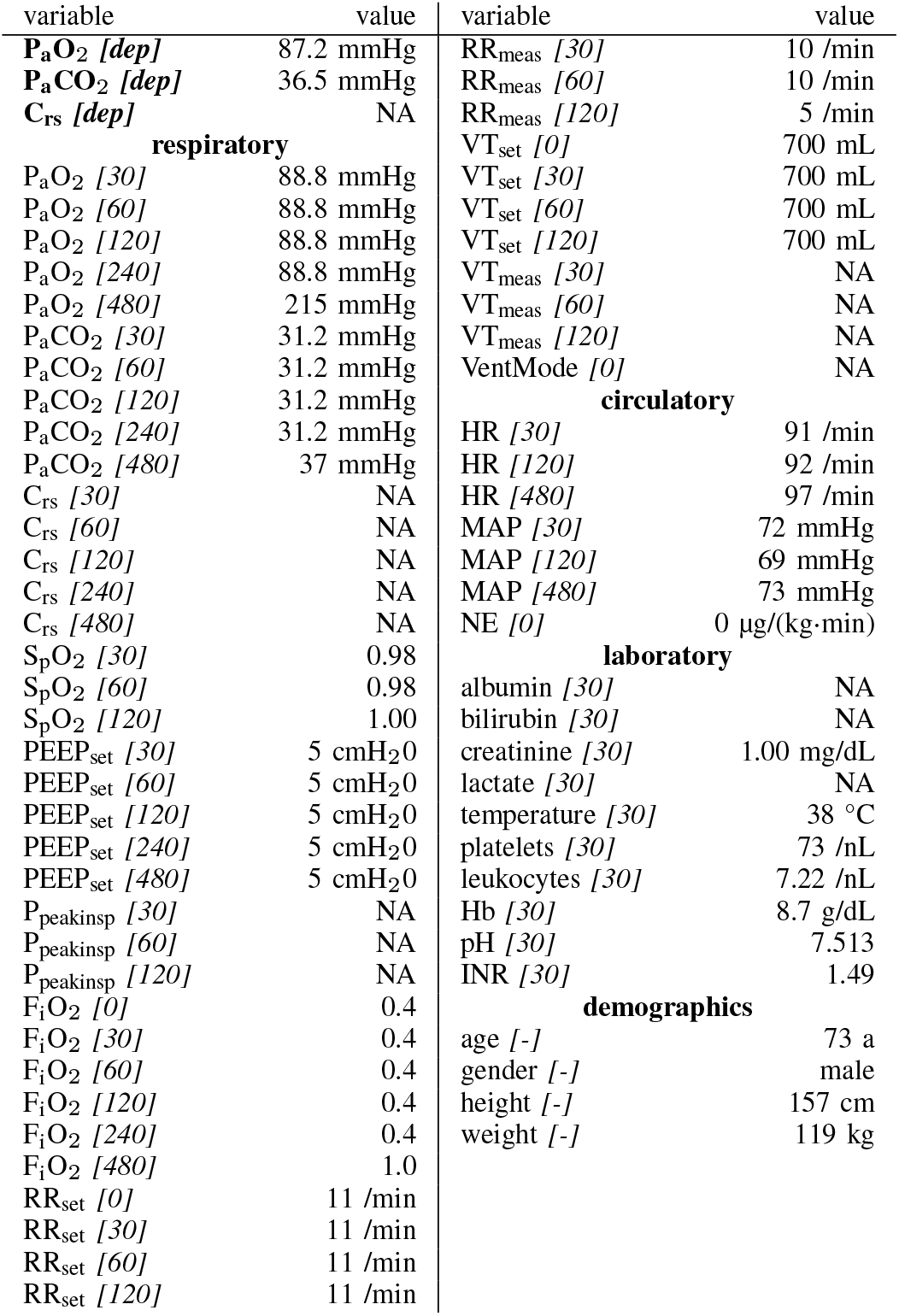
Example of a single sample for the *30 min blinded* setup, totalling 71 items per event. P_a_O_2_: arterial oxygen partial pressure, P_a_CO_2_: arterial carbon dioxide partial pressure, C_rs_: respiratory system compliance, S_p_O_2_: peripheral oxygen saturation (oximetry), PEEP: positive end-expiratory pressure, P_peakinsp_: peak inspiratory pressure, F_i_O_2_: inspired oxygen fraction, RR: respiratory rate, VT: tidal volume, HR: heart rate, MAP: mean arterial pressure, NE: norepinephrine rate, Hb: haemoglobin, INR: International Normalised Ratio; subscript *set*/*meas*: set/measured, numbers in square brackets indicate the time in minutes up to which the variable was blinded, *[dep]*: dependent variable, NA: not available.

In this paper, we present two similar setups, representing two clinical use cases. In the *all data* setup, the model predicts one or more target variables at a certain point in time, using all data that is available up to that time. This aims to help clinicians with real-time information instead of them having to wait for the results of a blood gas analysis or other measurement. Figure 1 illustrates how we selected and preprocessed measurements for the *all data* setup. The *30 min blinded* setup is identical to *all data* except that all measured variables (i.e. variables that are not set) in the last 30 minutes are removed. This corresponds to a prediction 30 to 60 minutes into the future. This model is used in the simulation setting which is explained below in section II-E.

**Fig. 1:**
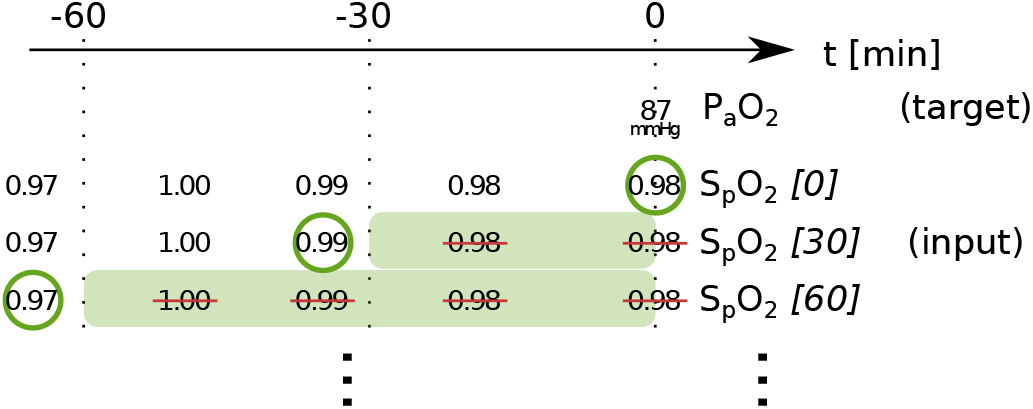
Preprocessing scheme for one observation of the target variable. Every row represents one item in the template, numbers in square brackets indicate the time in minutes up to which the variable was blinded, also represented by the shaded blocks indicating intervals that were discarded before selecting measurements; circles indicate selected measurements for every item.

From the MIMIC-III/eICU dataset, we included a total of 205736/133234, 205737/131800 and 207956/170851 individual observations for P_a_O_2_, P_a_CO_2_ and respiratory system compliance C_rs_, respectively.

### C. Model selection, training, and testing

The deep learning model uses a fully connected artificial neural network with 2 layers with 1024 and 512 nodes. Categorial variables were represented using embeddings of sizes 3 to 7. Settings for the training were as follows: Learning rate 2·10^−3^ with Adam optimiser, 20 epochs. For regularisation we used a weight decay of 0.1 and a dropout of 0.04, 0.05 and 0.5 for the embeddings and the first and second fully connected layers, respectively. We did not perform any hyperparameter tuning on test data.

With the used target variables, we care for relative rather than absolute errors. Therefore we opted for mean squared logarithmic error (MSLE) as our loss function. We trained the model for every target variable individually as well as together in the sense of multi-task learning [14]. In the latter case, the mean of the individual loss functions for each task was used as loss function. We used the MIMIC-III dataset as training and test set and evaluated our models’ performance independently on the eICU dataset. Implementation of the deep learning models was done with pytorch and the fast.ai framework [15]. We also trained a random forest model with the same data as a reference (50 trees, no maximum depth, loss function MSLE).

As metrics we report the mean absolute error (MAE) and the mean absolute percentage error (MAPE) of predictions for all target variables in the test set. To gauge performance transfer, we report performance on the held-back MIMIC-III test set along with the eICU test set.

### D. Interpretability

We estimated the feature importance of the input variables with a permutation approach [16]. To do this, the values for all single input variables were permuted across every observation in the batch in turn. The resulting difference in loss compared to the unpermuted data reflects the importance of any single input variable.

### E. Simulation

Based on the *30 min blinded* setup, we can make predictions about the expected effects of PEEP changes by presenting the model input data where the latest set PEEP value has been manipulated. As stated above, we carefully removed all measured variables for the time frame in question in order to blind the model from the real effects of the set PEEP that might have been altered by the manipulation.

## III. Results

Table II shows the results of both setups. We compared three computational settings: random forest (reference), individual models for all target variables and one multi-tasking model for all target variables.

**TABLE II:** Performance of models for both setups. Best (lowest) metrics highlighted. P_a_O_2_: arterial oxygen partial pressure, P_a_CO_2_: arterial carbon dioxide partial pressure, C_rs_: respiratory system compliance, MAE: mean absolute error, MAPE: mean absolute percentage error.

|  | random forest |  |  |  | neural network, individual models |  |  |  | neural network, multi-task |  |  |  |
| --- | --- | --- | --- | --- | --- | --- | --- | --- | --- | --- | --- | --- |
|  | MIMIC-III test set |  | eICU test set |  | MIMIC-III test set |  | eICU test set |  | MIMIC-III test set |  | eICU test set |  |
|  | MAE | MAPE | MAE | MAPE | MAE | MAPE | MAE | MAPE | MAE | MAPE | MAE | MAPE |
| *- all data -* |  |  |  |  |  |  |  |  |  |  |  |  |
| P<sub>a</sub>O<sub>2</sub> [mmHg] | **27.15** | 21.76% | 43.80 | **32.16%** | 28.66 | 22.49% | 46.98 | 42.56% | 27.16 | **21.11%** | **41.81** | 38.74% |
| P<sub>a</sub>CO<sub>2</sub> [mmHg] | 4.496 | 10.66% | 6.688 | 15.61% | 4.498 | 10.55% | 7.644 | 18.66% | **4.264** | **10.00%** | **5.492** | **13.27%** |
| C<sub>rs</sub> [mL/cmH<sub>2</sub>O] | 5.029 | 16.82% | 10.48 | 31.67% | 5.155 | 17.09% | 12.14 | 28.26% | **3.201** | **11.20%** | **9.195** | **22.54%** |
| *- 30 min blinded -* |  |  |  |  |  |  |  |  |  |  |  |  |
| P<sub>a</sub>O<sub>2</sub> [mmHg] | 28.16 | 22.67% | 44.68 | **33.12%** | 29.44 | 23.39% | 46.60 | 42.28% | **28.09** | **22.20%** | **43.04** | 41.17% |
| P<sub>a</sub>CO<sub>2</sub> [mmHg] | 4.547 | 10.79% | 6.804 | 15.83% | 4.557 | 10.68% | 7.509 | 18.26% | **4.338** | **10.15%** | **5.452** | **12.77%** |
| C<sub>rs</sub> [mL/cmH<sub>2</sub>O] | 5.059 | 17.07% | 10.17 | 29.71% | 5.169 | 17.21% | 11.92 | 28.25% | **3.240** | **11.33%** | **8.957** | **22.47%** |

Using the permutation approach, we calculated feature importance information for our best model resulting in table III.

**TABLE III:** Top 10 most important features ranked by permutation importance for different target variables (multi-task model for the *30 min blinded* setup); P_a_O_2_: arterial oxygen partial pressure, P_a_CO_2_: arterial carbon dioxide partial pressure, C_rs_: respiratory system compliance, F_i_O_2_: inspired oxygen fraction, PEEP: positive end-expiratory pressure, RR: respiratory rate, VentMode: ventilator mode, S_p_O_2_: peripheral oxygen saturation (oximetry), VT: tidal volume, P_peakinsp_: peak inspiratory pressure; subscript *set*/*meas*: set/measured, numbers in square brackets indicate the time in minutes up to which the variable was blinded.

| Top | P<sub>a</sub>O<sub>2</sub> | P<sub>a</sub>CO<sub>2</sub> | C<sub>rs</sub> |
| --- | --- | --- | --- |
| 1 | F<sub>i</sub>O<sub>2</sub> [0] | P<sub>a</sub>CO<sub>2</sub> [30] | C<sub>rs</sub> [30] |
| 2 | P<sub>a</sub>O<sub>2</sub> [30] | VT<sub>set</sub> [0] | VT<sub>set</sub> [0] |
| 3 | F<sub>i</sub>O<sub>2</sub> [30] | VentMode [0] | P<sub>peakinsp</sub> [30] |
| 4 | S<sub>p</sub>O<sub>2</sub> [30] | RR<sub>set</sub> [30] | C<sub>rs</sub> [480] |
| 5 | F<sub>i</sub>O<sub>2</sub> [120] | RR<sub>set</sub> [120] | RR<sub>set</sub> [0] |
| 6 | F<sub>i</sub>O<sub>2</sub> [480] | P<sub>a</sub>CO<sub>2</sub> [480] | VentMode [0] |
| 7 | PEEP<sub>set</sub> [30] | F<sub>i</sub>O<sub>2</sub> [0] | PEEP<sub>set</sub> [30] |
| 8 | RR<sub>set</sub> [0] | pH [30] | C<sub>rs</sub> [60] |
| 9 | VentMode [0] | P<sub>a</sub>CO<sub>2</sub> [60] | C<sub>rs</sub> [240] |
| 10 | S<sub>p</sub>O<sub>2</sub> [120] | VT<sub>meas</sub> [30] | RR<sub>set</sub> [120] |
| ... |  |  |  |
| 26 |  | PEEP<sub>set</sub> [30] |  |

Figure 2 shows simulations for a range of possible PEEP changes and their effect on the three target variables for two example inputs. Predictions for the three target variables are displayed in different colours over a shared X-axis, indicating the simulated PEEP. We see that the predictions and target values are in good accordance in these examples. In example **a** (patient from table I) P_a_O_2_ increases considerably with increasing PEEP while C_rs_ and P_a_CO_2_ remain mostly unchanged. Note that there are multiple missing input values and no ground truth measurement for C_rs_ in this example. In example **b**, increasing the PEEP is not expected to improve oxygenation.

**Fig. 2:**
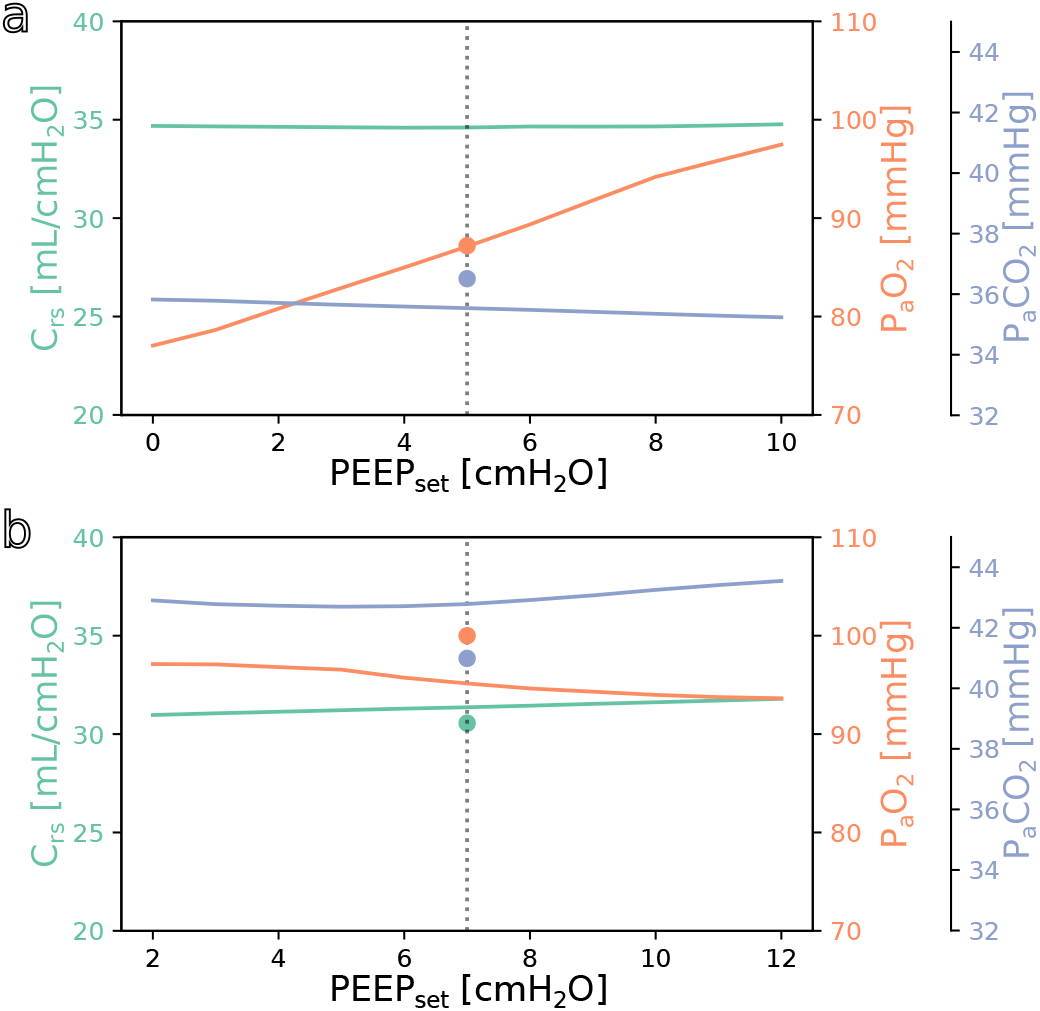
**a**: simulation for the above example with the values from table I as the only inputs. The vertical dotted line indicates the last set PEEP value. Circles denote the respective ground truth. **b**: same but for a different patient.

## IV. Discussion

In this paper we presented a deep learning based method to predict relevant success parameters of ventilation. With a similar model, simulations of PEEP changes and their effect on the success parameters can be explored.

We decided to use the mentioned input variables for this task with two goals in mind: on the one side, we included some measurements that provide information about the general state of health of the patient (mostly demographics and some variables also used in the SOFA score), on the other side we included measurements of common ventilator settings and measurements. With these, we were in parts limited by what values were available in both the MIMIC-III and the eICU databases. For our target variables we chose P_a_O_2_ and P_a_CO_2_, as these are the main indicators of successful gas exchange. Aside from the use as target variables, the oxygenation response to a PEEP change might also predict mortality [17], in this setting possibly even without actually changing the PEEP. We also included respiratory system compliance as one of the oldest indicators of “best PEEP” [18]. This parameter also helps to find ventilator settings with a low driving pressure and possibly a low transpulmonary pressure which might mitigate ventilator associated lung injury [19]. The time it takes for different variables to reach steady states after a PEEP change ranges from 5 to over 60 minutes and depends on the variables in question and on the direction of the PEEP step [20]. We chose 30 minutes as our prediction horizon as a compromise between the quantity of expected changes and the uncertainty and thus difficulty of the prediction task. Because of the binning of the variables, this choice is equivalent to a prediction 30 to 60 minutes into the future.

Methods for clinical decision support become more and more common. The strength of this tool is that it offers a very high degree of individualisation while at the same time being relatively easy to apply. Once the digital infrastructure is set up, the model takes a small number of routine measurements and outputs results - there is no need for additional material, measurements, interventions or manoeuvres. This simplicity is also true for the model itself, which not only makes it easier to understand and explain to clinical users, it also makes it possible to train the model in under 5 minutes on a single consumer graphics processing unit (GPU). This means that the model can easily be trained or fine-tuned regularly or when circumstances change, like during the beginning of a pandemic. Sometimes clinical decision support systems seem to take the power out of the clinician’s hand because they learn to rely on a proposed decision. The advantage of the solution presented here is that the clinicians still can and have to balance the expected effects on the different target variables and make their own decisions. We therefore refrain from offering concrete rules as to how one should react to certain model or simulation outputs.

With mean absolute percentage errors of 22%, 10% and 11% for the three tasks on the MIMIC-III test set, the model appears to reach a clinically useful performance level, especially for P_a_CO_2_ and C_rs_. To determine the caretaker acceptance and the perceived and actual benefit for patient care, a clinical study would be needed. When comparing the results of the *all data* and *30 min blinded* setup, the differences in prediction error are surprisingly small. Discarding 30 minutes of measurements does not seem to increase the difficulty of the task to a relevant degree. This may be in part explained by the fact that all target variables tend to change slowly, leaving the last known value as a very good guess. Another reason might be the high variability of the measurement of the ground truth, which for P_a_O_2_ and P_a_CO_2_ is subject to numerous preanalytical and (allowable) analytical errors [21], [22]. The fact that models were tested on a separate dataset from a different source and still performed well shows its generalisation capacity. The performance gap between the MIMIC-III test set (held separate from the training set) and the eICU test set is probably due to structural differences between the databases, their collection and the underlying treatments. We are confident that the real-world performance after fine-tuning with local data will be closer to the MIMIC-III test set performance. We reached our best result using the multi-tasking neural network. It outperforms both the random forest and the individual neural networks except for the MAPE metrics for P_a_O_2_ in the eICU test set in both setups and the MAE metric for P_a_O_2_ in the MIMIC test set in the *all data* setup, where the random forest performed better. The results clearly show the advantage of the multi-tasking model over the individual models. This is a known phenomenon for related tasks where the model may profit from the information gained from one task in another task [14]. Additionally, multi-tasking models simplify training and inference because one has to train, evaluate and store only one model. We chose the mean of the individual losses as our multi-tasking loss because of simplicity and because the individual losses were sufficiently similar in size as not to overpower one another.

Our analysis of feature importance shows that for all tasks high importance is attributed to features that seem clinically reasonable. Specifically, most important features naturally include the last known measurement of the variable to be predicted and variables that are closely correlated. For P_a_O_2_ these include inspired oxygen fraction and oximetric peripheral oxygen saturation, for P_a_CO_2_ these are the set tidal volume and set respiratory rate as well as pH, and for C_rs_ the set tidal volume and peak inspiratory pressure. For all target variables, the currently set ventilator mode occupies a top 10 position, showing that the model gathers valuable information from this categorial variable’s embeddings. The last known set PEEP occurs at positions 7, 26 and 7, respectively. Only the reason for the occurrence of the set respiratory rate (for P_a_O_2_ and C_rs_) and inspired oxygen fraction (for P_a_CO_2_) are not immediately obvious. It is important to note that this permutation based approach to determine feature importance should only be seen as a very rough peek under the model’s hood as it is ignorant of the complex interactions of multiple input variables because it considers every variable independently [23]. All in all, these results can be seen as a reason to have confidence in the decision process of the proposed model. This kind of interpretability is of major importance for the acceptance of machine learning solutions by clinicians and the population in general. Similar to the simulation approach used in this work, it would also be possible to better understand the model’s decision for a single case by manipulating or obscuring individual input variables.

From the simulation example (figure 2) we get an impression of how this tool might be used practically in clinical care. In the two shown cases, one has an expected increase in P_a_O_2_ in response to an increased PEEP while the other does not. This kind of graph might be useful for a clinician who is trying to optimise oxygenation or carbon dioxide elimination, especially in the short term.

A possible shortcoming of the simulation design we applied here is the risk of information leakage in the training and testing phases. Even if the model is blinded for all measured variables within the prediction horizon, it has still access to the set variables from this time period, which may be a clinician’s reaction to a measurement. However, this situation will not affect the model performance as no wrong information is introduced. As already mentioned above, the model succeeded to transfer knowledge from one US database to another. It could be more challenging, however, to transfer to the situation in Europe, for example because pressure controlled ventilation is much more common there. Additionally, medical knowledge has a short half-time for example as patient demographics change or new treatment options arise. What has been accurate for the MIMIC-III dataset recorded between 2001 and 2012 may be inaccurate in 2021. This makes fine-tuning on recent local data even more important. One big problem we encountered, as one does in many machine learning applications, was the quality of the data. While the MIMIC-III database is often praised for its quality, these large databases still suffer from missing and corrupt data or incomplete documentation. This becomes especially evident when moving across different databases. Finally, due to the chosen approach in this work, it is not easily possible to include long-term clinical endpoints like mortality into the prediction because we use a fixed prediction horizon. For these endpoints the much more complicated reinforcement learning approach would be necessary [11].

To be able to use the developed model in Europe, we plan to continue training the model on data from local ICUs. A possible next step would be to include more ventilator settings into the simulation part, eventually making it possible to try out different ventilator modes, pressures and timings without even having to bother (and possibly harm) the patient. The relatively simple process and model design we applied to a ventilation problem here would also be suited for a wide range of applications outside of ventilation therapy, for example the prediction of drug concentrations, regulation of coagulation, fluid balance or circulation. For all these applications, clinicians currently rely mostly on experience or rules of thumb and would definitely benefit from a reliable and objective prediction.

## V. Conclusion

Deep learning for tabular data is a useful approach for this clinically relevant problem of ventilation optimisation. It delivers reliable results and comes with no extra cost once the digital infrastructure is in place. There is a large potential for this tabular approach for numerous other applications in the ICU.

## Data Availability

Datasets are used from the collection of eICU Collaborative Research Database and Medical Information Mart for Intensive Care III.
We passed the Protecting Human Research Participants exam and obtained permission to access the dataset. Access to the data (via physionet) was granted after registration on Oct. 23, 2019, 3:47 p.m. Necessary Agreements were signed:
Nov. 7, 2019 PhysioNet Credentialed Health Data Use Agreement 1.5.0 eICU Collaborative Research Database (v2.0);
Oct. 24, 2019 PhysioNet Credentialed Health Data Use Agreement 1.5.0 MIMIC-III Clinical Database (v1.4)

https://eicu-crd.mit.edu/

https://mimic.physionet.org/

